# Towards accesible brain-tumor classification at the point of care: Nanopore methylation sequencing from Formalin-Fixed Paraffin-Embedded (FFPE) pathology samples

**DOI:** 10.1101/2024.05.02.24306400

**Authors:** Galina Feinberg-Gorenshtein, Assaf Grunwald, Carlo Vermeulen, Nurit Gal Mark, Lena Shinderman Maman, Keren Shihrur, Michal Hameiri-Grossman, Orly Michaeli, Suzanna Fichman, Abraham Natan, Tali Siegal, Shlomit Yust-Katz, Hanna Weiss, Adva Levi-Barda, Osnat Konen, Amir Kershenovich, Jeroen de Ridder, Helen Toledano, Yehudit Birger, Shai Izraeli, Yuval Ebenstein

## Abstract

Oxford Nanopore Technology (ONT) based methylation sequencing is increasingly recognized for its rapid and accurate classification of brain tumors. A process that is crucial for optimal patient treatment. However, widespread clinical utility is currently limited by the need for fresh-frozen biopsies and not the standard-of-care formalin-fixed, paraffin-embedded (FFPE) samples. Our study explores the impact of FFPE on DNA methylation and presents a developed and validated protocol for ONT-based FFPE tumor classification. We present a practical solution for precise brain tumor diagnoses in routine clinical settings and facilitating timely treatment decisions at the point of care and without interfering with operating room standards.

---

Accurate categorization of tumor types and subtypes is crucial for facilitating comprehensive precise patient treatment ^1–3^. For instance, the World Health Organization (WHO) classification of central nervous system (CNS) tumors encompasses over 10 major tumor classes, each comprising numerous subclasses, totalling more than 100 distinct entities that necessitate diverse treatments and are associated with different prognosis and clinical courses^4^. These tumor entities frequently exhibit resemblances in morphological, spatial, and genetic characteristics^5–7^, rendering their differentiation challenging. Moreover, neuropathologists may provide varying interpretations of histopathological results, adding subjectivity to the process^8^.

Epigenetics, on the other hand and particularly DNA methylation—has proven to be a robust and stable tool for accurately distinguishing the vast majority of these tumor subtypes^2^ hence methylation based classification has been incorporated into the 2021 WHO classification of CNS tumors^4^. Consequently, the examination of tumor methylation patterns has recently become a part of clinical diagnostic procedures, with methylation-based classifiers already available for CNS tumors and sarcomas, and others under development for different tumor types^9^.

Yet, the incorporation of DNA methylation into the diagnostic workup of brain tumors has proved to be challenging. The “gold standard” method to assess these methylation patterns is DNA hybridization arrays, such as the Infinium MethylationEPIC array^10^ comes with significant drawbacks, including high expenses, long turnover times, high amount of required starting material (minimum 500ng DNA, preferably 1ug DNA), need to accumulate several samples to get results and the need for highly trained personnel, incongruent with the short timeframes often mandated by clinical needs.

To address these challenges, a number of recent studies have successfully employed Oxford Nanopore Technology (ONT), known for its rapid and relatively straightforward epigenetic sequencing, to obtain the necessary methylation patterns for classification ^11–14^. Subsequently, several healthcare centres have begun integrating this technology into patient diagnosis^15^.

Nevertheless, the widespread adoption of ONT for tumor classification and other biopsy-originated DNA analyses faces a major obstacle. Unlike hybridization arrays, ONT requires concentrated DNA samples of high purity and high molecular weight such as purified from freshly frozen (FF) biopsy samples. Currently, most health centres can only provide Formalin-Fixed Paraffin-Embedded (FFPE) samples due transport and storage limitations^8,16^. Additionally, the unknown tumor fraction in FF samples poses a degree of uncertainty, since a low tumor fraction in the homogonized sample limits classification^17^. In FFPE samples, histopathological staining can be performed, so the tumor fraction is not only known but can be selectively enriched for analysis^18^. An example is presented in figure 1; in this case, histopathological staining of low tumor-fraction sample, allowed to extract DNA only from regions with relatively high tumor-fraction of 20-30% and to perform downstream analysis on these regions only.

**Figure 1:**
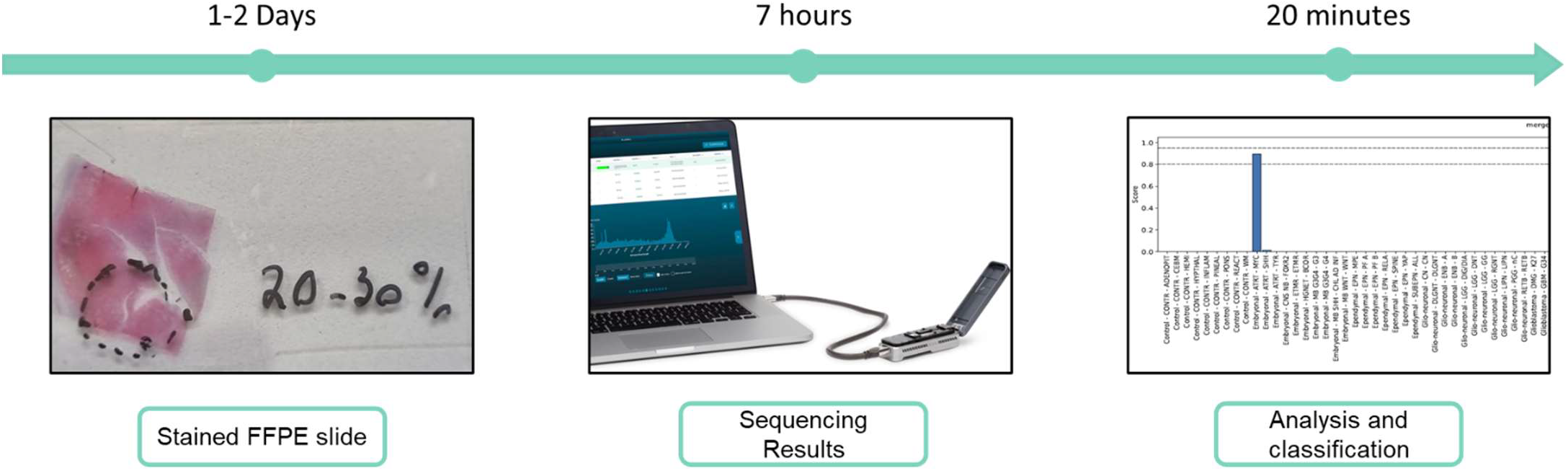
Schematic representation of the typical timeline and steps of our suggested protocol – from biopsy to classification: Left: Generation of FFPE slides with histochemical staining typically takes 1-2 days. Middle: Our adjusted protocol for DNA purification from histological slides takes ∼4 hours followed by roughly 90 minutes for adjusted ONT DNA library preparation and ∼120 minutes of sequencing are required to gather a sufficient amount of data for classification (∼7 hours all together). Right: “Sturgeon” classification of sequencing data takes ∼20 minutes on a local computer.

Given the clinical widespread ability to analyse FFPE samples rather than FF specimens, we validated an FFPE procedure that does not disrupt the accurate classification achieved through nanopore methylation sequencing. Subsequently, we developed a protocol for methylation sequencing and classification of FFPE samples. We were able to successfully classify DNA derived from FFPE samples, enabling the precise classification of brain tumors within 30-55 hours, synchronized with standard of care (Figure 1).

We first wished to verify that the FFPE process does not hamper DNA methylation-based classification. Tumor samples were collected from six patients and processed as both FFPE and FF. Samples were subjected to DNA extraction and methylation sequencing (see methods for adjusted FFPE protocol). Despite clear methylation altreations due to the FFPE treatment (SI Figure 1), all samples were classified correctly under both storage conditions. To test whether FFPE generates sequence-biased methylation alterations, we performed the following analysis: All CpGs in the genome were grouped according to their 4-base sequence context (ACGA, ACGC etc. 16 combinations in total, Figure 2), next the average methylation levels were calculated for each group. We compared methylation levels of these sequence groups in the data generated for the six sample pairs: In two sample, stored in formalin prior to block generation for 1-2 days, no significant difference was found in methylation levels between FF and FFPE treatment (p-values of 0.99 and 0.07, SI table 1). For the other four pairs, stored in formalin prior to block generation for 1-2 days, we conducted bootstrapped confidence interval analysis, examining the difference in methylation means for each of these sites within each pair (see SI for details). These analyses indicate that although methylation is decreased in FFPE samples, the decrease is not sequence specific (Figure 2 and SI table 1), thus the relative methylation levels and patterns remain, allowing accurate tumor classification of FFPE samples.

**Figure 2:**
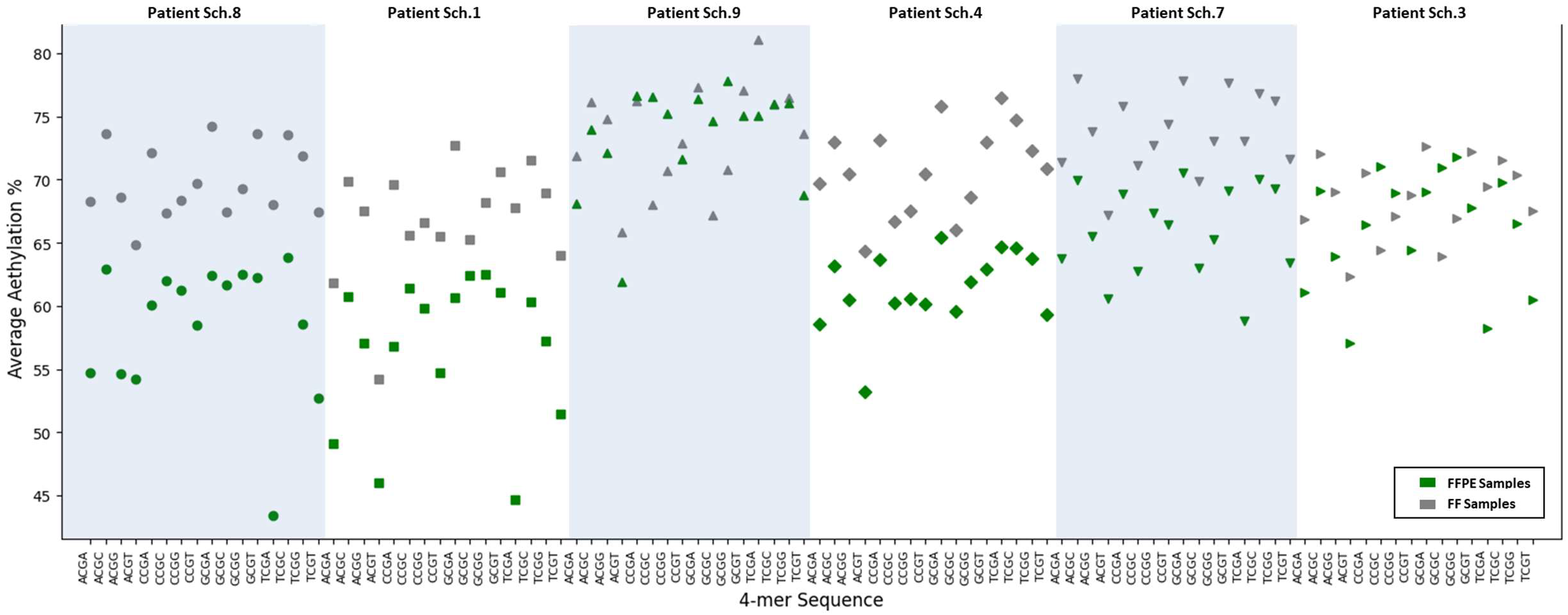
comparison of methylation levels between FF and FFPE for six biopsy samples that were processed by both types of preservation methods. The average methylation level for all possible combinations of 4-base sequences containing CpG at positions 2 and 3 (as indicted on the X-axis for each patient) are depicted for each patient, with different patients presented individually along the X-axis. FF data are shown in grey, and FFPE data are depicted in green. The image clearly indicates that patients Sch.9 and Sch.3 exhibit similar methylation levels in both FF and FFPE samples. In contrast, for the remaining patients, FF samples demonstrate higher methylation levels.

Next, given the low amounts of input material available from many FFPE blocks we wished to assess the limiting DNA amount for classification. Purified DNA form previously classified FFPE samples was devided into 4 subgroups with varying amounts of extracted DNA, ranging for 25ng to 1000ng. We performed methylation sequencing and classification for each subgroup (see methods for adjusted FFPE protocol), obtaining correct classification even for the lowest input amount of 25ng DNA (SI table 2). We note that this amount is 5 times lower than the minimal amount recommended by ONT for sequencing and 40 times lower than the amount required for DNA array analysis.

To validate the clinical applicability of FFPE samples for methylation-based classification, we retrospectively sequenced 13 DNA samples purified from FFPE CNS tumor tissues of pediatric patients using ONT methylation sequencing. The classification was performed using the Sturgeon tool which provides a confidence score for the classified entity^11^. This cohort was previously classified through various gold-standard clinical methods (SI Table 3).

Remarkably, all classifications were consistent with those provided by previous methodologies. 38% of the samples (5 samples) were classified with a confidence score of 0.95 and above, another 38% of the samples (5 samples) were classified with a confidence score of 0.95-0.9, and the remaining 23% of samples (3 samples) were classified with a confidence score below 0.9, as detailed in SI Table 3.

To underscore the utility of our approach for accurate diagnosis, enabling potentially life-saving treatment decisions, we present three case studies:

Case A (Patient Sch.3) involves a 0-5-year-old male presented with right hemiparesis. An MRI revealed a large left parieto-temporal tumor with a large metastasis in the suprasellar region behind the optic chiasm and suspected leptomeningeal spread (Figure SI.4.a). Initially treated with upfront chemotherapy without biopsy, the patient experienced clinical and radiographic deterioration. Following, a gross total resection of the temporal lesion was performed, initially suspected by the surgeon as a low-grade glioma. Initial histopathological analysis of a FFPE biopsis failed to determine classification. However the ONT sequencing protocol confidently classified this tumor as “Meningioma”. Following this finding repeated pathological analysis of the old and new specimens were compatible with meningioma.

4 days post-op the patient developed sudden onset blindness. Our meningioma classification has guided the following paitent treatment: to avoid radiation in such a young child, and since meningioms are insensitivie to chemotherapy, it was decided to perform debulking of the suprasellar tumor and relieve pressure on the chiasm with restoration of vision.

Case B. (Patient Sch.1) involves a 6-10-year-old female presented with vomiting and left facial nerve palsy. An MRI revealed a heterogeneous mass in the left cerebellopontine angle CPA (Figure SI.4.b) the tumor underwent subtotal resection and pathology strongly suggested a diagnosis of Atypical Rhabdoid Tumor “ATRT”, however, staining for INI1 (SMARCB1) showed a mosaic pattern (Figure SI.4.c), resulting in unconfirmed diagnosis. Chromosomal microarray analysis (CMA) ( Cytoscan HD, Thermo Fisher Scientific), identified only mono-allelic loss of the SMARCB1 gene and an NGS sequencing panel, (Oncomine Childhood Cancer Research Assay; Thermo Fisher Scientific) was also unable to identify pathogenic or likely pathogenic variants in thecoding regions of SMARCB1 and SMARCA4 genes. Nevertheless, our sequencing protocol on FFPE tissue allowed to classify the tumor as “ATRT, MYC subgroup”, resulting in initation of appropriate chemotherapy. This diagnosis was subsequently reconfirmed a few weeks later by Infinium MethylationEPIC array (Brain tumor classifier; Version:11b4).

Case C. (Patient Sch.9) involves a 0-5-female from whom MRI revealed a spinal cord lesion extending from L2-S2 (Figure 3.a). Neuroblastoma was suspected based on clinical and imaging findings. CT guided biopsy was performed (Figure 3.b), however, due to the challenging tumor location and the low weight of the infant, only a small FFPE tissue block was available. Due to the low tissue amounts and poor DNA quility, pathology and an NGS panel were non-informative, and diagnosis couldn’t be determined. However, our sequencing protocol allowed to classify the low FFPE derived DNA amounts (120ngr) as “ATRT, MYC subgroup”. Age adjusted chemotherapy according to the EuRhab protocol was initiated. A subsequent open biopsy confirmed the diagnosis: immunohistochemistry was negative (absent staining) for INI1, and CMA ( Cytoscan HD, Thermo Fisher Scientific) and an NGS panel (Oncomine Childhood Cancer Research Assay; Thermo Fisher Scientific), identified a bi-allelic deletion of the SMARCB1 gene, associated with this tumor class^19^.

**Figure 3:**
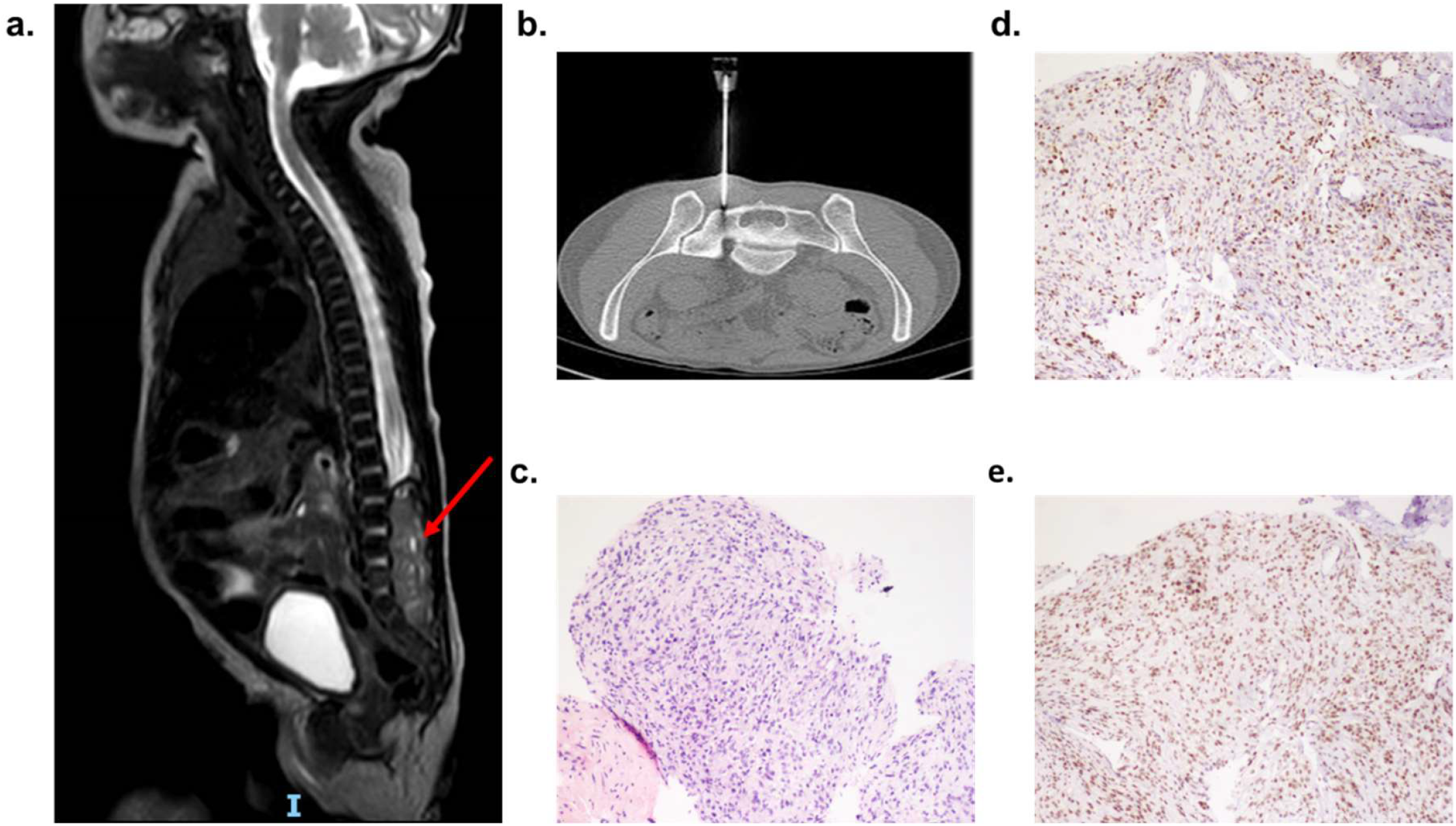
Patient Sch.9: **a**. MRI shows tumor filling the spinal canal at the level of L2-S2, T2 sagittal view. Tumor showed heterogeneous enhancement post contrast. **b**. CT guided needle biopsy provided only 120 ng of DNA of poor quality. **c**. H&E staining of Atypical teratoid rhabdoid tumor, WHO grade 4. **d**. SMARCB1 (INI1) immunohistochemistry staining showing nuclear loss of expression in tumor cells and retained expression in endothelial cells. **e**. BRG1 immunohistochemistry staining.

In the examples above, epigenetic ONT sequencing provided early, rapid, and accurate classification that enabled prompt management decisions and initiation of appropriate and timely treatment. Our protocol for FFPE analysis will enable the adoption of the ONT diagnostic tool for routine clinical use in health centers, where most of the time, only FFPE samples are available. This approach for CNS tumor classification can be extrapolated for utilization in other genetic or epigenetic testing scenarios.

## Methods

### Sample Processing and DNA Extraction

DNA library preparation for ONT sequencing requires high molecular weight DNA at high concentrations and at high degree of purity. To address DNA length and purity we made several modifications to the DNA extraction and library preparation protocols:

DNA extraction from FFPE samples was performed after tumor content evaluation using the QIAamp DNA FFPE tissue kit (Qiagene, Hilden, Germany) or the RecoverAll Multi-Sample RNA/DNA Kit (utilizing DNA only, Invitrogen, Thermo Fisher). To facilitate ONT sequencing, specific adjustments were implemented: Instead of the prescribed paraffin removal using Xylene in the protocol, tissue sections from 7-17 slides were accumulated in a 1.5mL tube containing 400 µL Digestion buffer Subsequently, the tubes underwent heating in a block (at 90°C for 3 minutes), followed by centrifugation 14,000g for 1 minute. After 1 minute on ice, the resulting paraffin ring was extracted. DNA from Fresh tumor tissue (FF) was extracted using spin columns (QIAamp DNA Micro Kit, Qiagen) according to the manufacturer’s protocol. The quantification of the isolated DNA was conducted using a Qubit Flex fluorometer with a 1x dsDNA HS Assay Kit (Invitrogen, Thermo Fisher). DNA purity was assessed using NanoDrop (Thermo Fisher Wilmington, DE, USA) at 260 and 280□nm.

H&E staining of one slide allowed for accurate scribing of tissue sections from slides and unnecessary paraffin removal. In cases of non-uniform distribution of tumor cells across the slide, H&E staining allowed pathologists to identify and indicate regions with high tumor fraction and to purify DNA from these regions only, increasing tumor fraction in the sequenced final sample.

This protocol was validated for FFPE blocks stored for up to 36 months.

### Library preparation and Nanopore Sequencing

FFPE samples were prepared for sequencing using Ligation Sequencing Kit V14 (SQK-LSK114, Oxford Nanopore Technologies, UK) with the following adjustments:

In ‘DNA repair and end prep’ section, incubation time was extended to 30 minutes at 20°C and 30 minutes at 65°C.

Bead-to-sample ratio was increased during purification to 180ul of beads in ‘DNA repair and end prep’ section and to 120ul of beads in ‘Adapter ligation and clean-up’ section. Ligation time was increased to 30min in ‘Adapter ligation and clean-up’ section and elution volume was reduced to 12ul. FF samples were prepared for sequencing using Rapid Sequencing Kit V14 (SQK-RAD114, Oxford Nanopore Technologies, UK) according to manufactor’s recommendations. Whole genome sequencing was performed on a MinION Mk1C device using R10.4.1 Flow cells (FLO-MIN114, Oxford Nanopore Technologies). For FFPE libraries minimum read length was set to 20 bp.

### Data processing and CNS classification

Basecalling of raw POD5 files was performed using the ONT proprietary software (Guppy v 6.4.6, Oxford Nanopore Technologies, UK). With the following model: “dna_r10.4.1_e8.2_400bps_modbases_5mc_cg_hac_mk1c.cfg”. Reads were then aligned to the T2T-CHM13 human reference genome using minimap2 v.2.24^20^. bam output files were than merged, sorted and indexed using samtools version1.16.1^21^. Merged and sorted bam were gave as an input to Sturgeon classifierr^11^ for CNS classification using default parameters. Analysis were performed a Linux operating system (Ubuntu 22.04.3) with Nvidia’s Getorce RTX 3060 GPU.

## Supporting information

Main SI file

## Data Availability

All data produced in the present study are available upon reasonable request to the authors

