## Supplementary material for "Towards accesible brain-tumor classification at the point of care: Nanopore methylation sequencing from Formalin-Fixed Paraffin-Embedded (FFPE) pathology samples": Main SI file

*Galina Feinberg-Gorenshtein^1^, *Assaf Grunwald^2,^, Carlo Vermeulen^3^ *, Nurit Gal Mark^1,5^, Lena Shinderman Maman^1^*, Keren Shihrur^1^, Michal Hameiri-Grossman^1^, Orly Michaeli^1^, Suzanna Fichman^6^, Abraham Natan^6^, *Tali Siegal^6^, Shlomit Yust-Katz^6^,* [*Hanna Weiss*](https://pubmed.ncbi.nlm.nih.gov/?term=Weiss+H&cauthor_id=17685476)*^6^, Adva Levi-Barda^6^,* Osnat Konen^7^, Amir  Kershenovich^8^, *Jeroen de Ridder^3^*, *Helen Toledano^1,4^*, Yehudit Birger^1,3,5^, Shai Izraeli^1,3,5^, and [Yuval Ebenstein](https://pubmed.ncbi.nlm.nih.gov/?term=Ebenstein+Y&cauthor_id=30485249)^2^

* Equal contribution

1. The Rina Zaizov Division of Pediatric Hematology-Oncology, Hemato-Oncology Laboratory, Schneider Children's Medical Center of Israel, Petach Tikva, Israel
2. Department of Physical Chemistry, School of Chemistry, Tel Aviv University, Tel Aviv, Israel
3. Center for Molecular Medicine, University Medical Center Utrecht, Utrecht, Holland.
4. Department of Human Molecular Genetics and Biochemistry, Faculty of Medicine, Tel Aviv University, Tel Aviv, Israel
5. Felsenstein Medical Research Center, Tel Aviv University, Israel
6. Department of Pathology, Beilinson Hospital Institute of Pathology, Petach Tikva, Israel
7. The Institute of Imaging, Schneider Children's Medical Center of Israel, Petach Tikva, Israel.
8. Department of Pediatric Neurosurgery, Schneider Children's Medical Center of Israel, Petach Tikva, Israel

**Limit of detection Assessment for Reliable CNS Classification**

To study sequencing and classification of low amounts of DNA, often associated with FFPE samples, and to determine the minimum DNA amount threshold for classification, we divided DNA purified from two FFPE CNS tumors, into 4 sub-samples of decreasing amounts (SITable 1.) and ran independent sequencing experiments with each.

In both DNA groups, a correct (concordant with classification made by other methods) and significant classification was achieved even for the lowest amount tested (25 ng, SI Table 1.). This amount is 5 times lower than the standard DNA amount in ONT experiments and allows diagnosis even from small pediatric biopsies.

| Paitent ID | DNA Quantity (ng) | DNA Quality (260/280) | DNA Quality (260/230) | No. of probes | Significance Score | Classification |
| --- | --- | --- | --- | --- | --- | --- |
| Sch.6 | 1000 | 1.8 | 2.08 | 953 | 0.93 | EPN-PF A |
| Sch.6 | 100 | 1.8 | 2.08 | 2347 | 0.99 | EPN-PF A |
| Sch.6 | 50 | 1.8 | 2.08 | 1932 | 0.99 | EPN-PF A |
| Sch.6 | 25 | 1.8 | 2.08 | 1749 | 0.93 | EPN-PF A |
| Sch.3 | 800 | 1.7 | 1.05 | 15647 | 0.97 | MNG |
| Sch.3 | 100 | 1.7 | 1.05 | 4071 | 0.97 | MNG |
| Sch.3 | 50 | 1.7 | 1.05 | 4667 | 0.97 | MNG |
| Sch.3 | 25 | 1.7 | 1.05 | 1430 | 0.96 | MNG |

***SI Table 1. determination of minimal DNA amount required for classification.***

*DNA from two patients was divided into 4 groups of decreasing DNA amounts, as detailed in the table. Independent sequencing experiments and classification were performed with each group. Classification results scores and number of probes used for classification (see methods) are detailed from each experimental group.*

**FFPE effect on methylation**

The generation of FFPE blocks involves the application of various chemicals to tissue, prompting concerns about potential chemical impacts on DNA, including changes in DNA modifications like 5mC. Despite prior studies dismissing this concern ^1,2^, and the routine use of FFPE-derived DNA for methylation analysis for clinical uses ^3,4^, we conducted a specific investigation into the impact of FFPE procedures on our experimental set.

To address this, we examined six patients who underwent both FF (fresh-frozen) and FFPE tissue biopsies. Initially, we assessed global 5mC levels using the Dorado basecaller by ONT (Kuśmirek, 2023). We observed a significant reduction in 5mC levels in FFPE samples compared to FF samples (in average 70.6% of CpG sites are methylated in FF samples and 63.7% for FFPE samples, p-value<0.05, paired t-test, SI Figure 1).

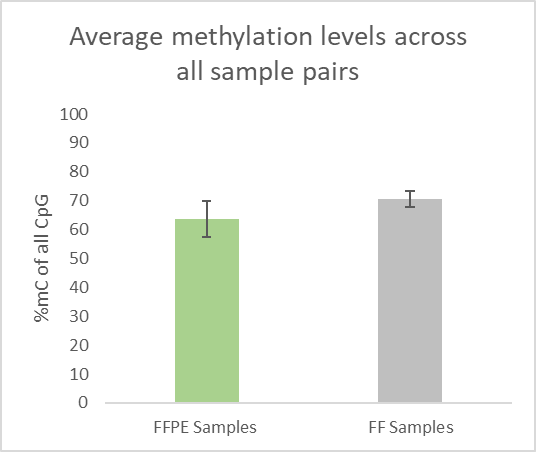

***SI Figure 1. Global reduction in 5mC level in FFPE samples***

*In the bar graph, average methylation values are presented for all CpG sites for six patients with paired biopsies, FF samples average is shown in the right bar (grey color) and FFPE in the left bar (green color) error bars show standard deviation values.*

Given these results and the critical need to ensure that FFPE does not alter modifications in a way that could impact tumor classification, we sought to understand the nature of the chemical effect of FFPE on DNA. Specifically, we determined whether this effect is due to a global stochastic process, where methylation levels change in similar extent along the DNA, or if FFPE mediates different changes on different CpG in a sequence specific manner. This is crucial because stochastic changes would preserve relative methylation patterns, enabling tumor classification. Conversely, a sequence-specific chemical process might bias relative methylation levels at certain sites, impeding classification.

To achieve this, we calculated average methylation levels for each of the 16 possible combinations of 4-mers containing CpG at positions 2 and 3 across the human genome (ACGA, ACGC, CCGA, etc., see Figure 1) for each of the 6 sample pairs (FF and FFPE, 12 in total) (Figure 1).

T-test analysis showed no significant difference in methylation levels for the different sites for two of the patients (SI table 1). For the remaining four patients, for whom the difference was significant (SI table 1) we assessed whether the difference between FF and FFPE methylation levels follows a stochastic or biased pattern. To address this, we conducted bootstrapped confidence interval analysis, examining the difference in methylation means for each of these sites within each pair (see below). A confidence interval containing zero indicates no bias towards specific sequence context. The results indicated a stochastic reduction in methylation levels for the different 4-mer sites in FFPE samples for all four patients (Figure 1. SI table 1).

| **Patient** | **2** | **3** | **4** | **5** | **6** | **7** |
| --- | --- | --- | --- | --- | --- | --- |
| **t-test** | 2.30674E-07 | 0.0700865 | 4.74496E-11 | 5.09718E-08 | 2.4048E-12 | 0.993936 |
| **Bootstrap Confidence Interval** | 8.1-12.6 | -0.03-4.8 | 7.1-8.9 | 9.4-13.8 | 8.5-10.3 | -2.2-2.0 |

***SI Table 2. Results of samples pairs statistical analysis***

*t-test and bootstrap confidence interval values for methylation values at different 4-mer context for all sample pairs. Patients 7&3 (highlighted in green) exhibited no significant difference between FF and FFPE samples. The other four patients (highlighted in blue) exhibited significant difference (p-value < 0.05) however their confidence interval does not contain zero indicating that the difference in methylation is stochastic and systematically reduced.*

Briefly these analyses were done as follows: pod5 files were basecalled using Dorado (version: 0.3.4, Oxford Nanopore Technologies, UK) with the “dna_r10.4.1_e8.2_400bps_hac@v4.2.0” model and “-modified-bases 5mCG_5hmCG” flag for modified bases calling (Oxford Nanopore Technologies, UK). Reads were then aligned to the hg38 human reference genome using minimap2 v.2.24^5^. bam output files were than merged, sorted and indexed using samtools version1.16.1^6^. These files were gave as an input to modkit (version: 0.1.12, Oxford Nanopore Technologies, UK) with the pileup command for extraction of methylation levels with specific CpG locations.

To extract locations of all possible 4-mers combinations with CpG at positions 2&3 in a bed format, an in-house python script was used. Following, the resulted bed files were intersected (using bedtools v2.30.0^7^) with the output files from the modkit pileup to generate a location and score file for each sample and 4-mer, methylation values were averaged for each 4-mer for each file. These values were used as input for t-test and bootstrap analysis which were performed using an in-house python script: briefly 1000 bootstrap iterations were performed where each time the paired mean difference was calculated. For each sample pair, the width and location of 95% confidence interval are presented in SI Table 2. When 0 is included within this interval the difference in the sample pair is systematic. Python scripts are added as Supporting Information.

Interestingly, the systematic difference is correlated not with the overall storage time but with the time the sample spent in formalin before the creation of the FFPE block: The more time the tissue spent in formalin a greater 5mC reduction effect is observed compared to FF samples (SI figure 2).

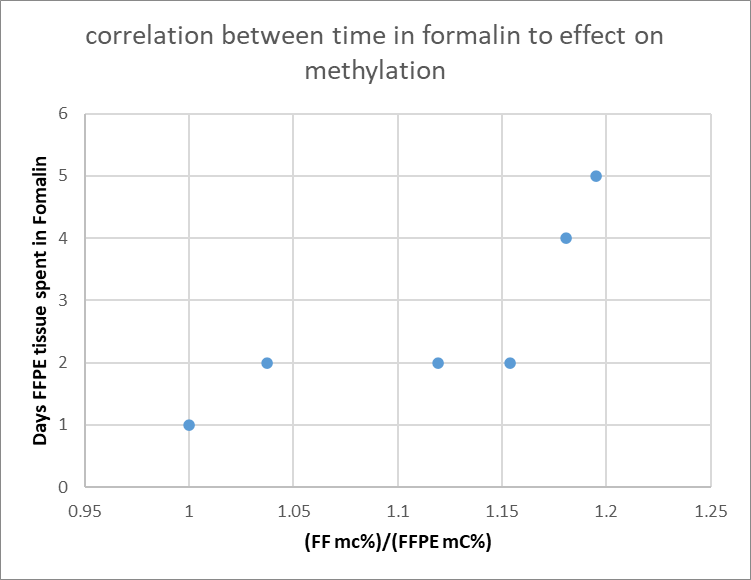

***SI Figure 2. Correlation between time in formalin and effect on methylation:*** *Days the tumor tissue spent in formalin prior to FFPE block creation are plotted against the decrease in methylation compared to the paired FF samples (the ratio between the average methylation in FF sample and average methylation in FFPE sample). It is clear that the longer the tissue spent in formalin the greater the reduction in methylation.*

For a deeper comprehension of how formalin affects DNA, a specialized study is necessary. Nonetheless, these findings imply that formalin acts as the agent impacting methylation before the creation of FFPE blocks (which involves tissue washing).

Finaly, we sought to explore effects on individual CpG sites in the genome. For this purpose, we computed methylation scores for each CpG site in all samples using modkit (version: 0.1.12, Oxford Nanopore Technologies, UK). The scatter plot presents the methylation level of each CpG in the FFPE sample (Y axis) and in the FF sample (X axis), showing a linear correlation within sample pairs (SIFigure 3). These analyses confirm that when FFPE affects DNA methylation, the effect is systematic across the genome in a way that preserves relative methylation levels, enabling tumour classification.

For this comparison we choose only CpG sites that had coverage of 5X or greater, using AWK and bedtools intersect command (version v2.30.0^7^). Linear fir to scatter plots was performed with python’s (version 3.10.7) numpy (numpy 1.32.2) polyfit option. Outlayers for scatter plot were detected by first calculating the inversion of the covariance matrix of both variables, than calculating the mahalanobis distance of each value in the matrix and finally only values with chi square distribution larger than 0.65 were highlighted (SI figure 3) all performed with python’s numpy package.

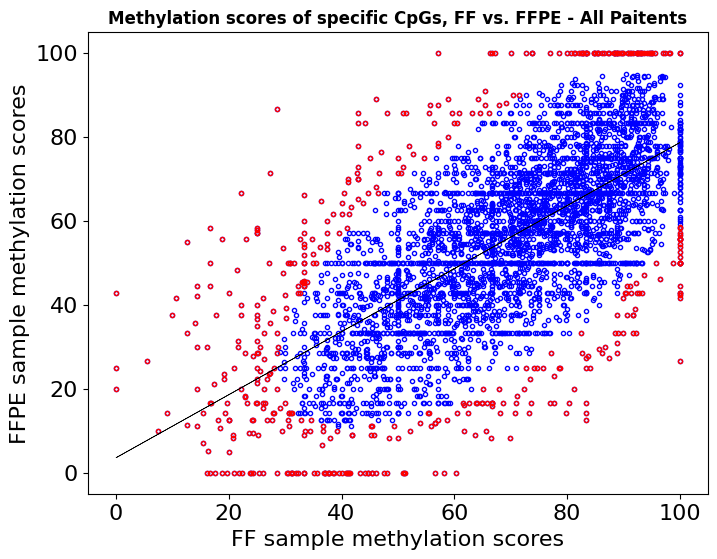

***SI Figure 3. Methylation values of individual CpG sites in FFPE vs***

***FF samples***

*Methylation values at specific CpGs of FF samples (X-axis) are plotted against these values of FFPE samples (Y-axis). Outliers are colored in red. This plot includes CpG values of all six patients with coverage > 5.*

**Sturgeon Classifier Analysis**

Sturgeon was trained on the Capper et al. 2018 dataset which was collected using the illumina 450K methylation array. Sturgeon is therefore limited to using these ~450.000 sites (probes) as input vectors. Sturgeon was developed to recognize any combination of the 450K array, and a depth of 1000-50000 sites. When an input vector containing the covered probes and their methylation status is provided, Sturgeon returns a confidence score for each of the tumor classes. A score over 0.95 is considered highly confident, and scores between 0.8 and 0.95 are considered somewhat reliable but should be treated with some caution. Scores below 0.8 are considered unreliable.

**Assay Validation**

To validate clinical applicability of DNA from FFPE samples for methylation-based classification, we retrospectively sequenced 13 DNA samples purified from FFPE CNS tumor tissues of pediatric patients using ONT sequencing (see methods for adjusted FFPE protocol). The classification was performed using the Sturgeon tool which provides a confidence score for the classified entity^8^. This cohort was previously classified through various methods, including Infinium HumanMethylation450, BeadChip EPIC array, Foundation One, standard histopathological examination, and other molecular tests (SI Table 3). The cohort was based on availble CNS padriatric FFPE samples obtained at diagnosis. All the availble samples with suffcient DNA amounts (above 25 ng) are reported in table 1.

A total of 19 tissue biopsies from 13 patients and the corresponding clinical data were obtained from the Pediatric Hematology-Oncology Department at the Schneider Children’s Medical Center of Israel. The study was conducted in accordance with the Declaration of Helsinki, under approval of The Rabin Medical Center (RMC) Institutional Review Board) IRB# 0012-08-RMC), led by Prof. Tur-Kaspa (Zeev Jabotinsky St 39, Petah Tikva, Israel, 4941492), and was carried out according to local guidelines and regulations. We obtained written or electronic informed consent by all participants or their legal representatives to take part in the present study and their consent to publish the results of this study., approval No. 920061517.

Despite differences in diagnosis and classification terminology between the Sturegon classifier and the pathology reports, we note that all classifications were found in agreement by qualified pathologists.

| **Patient’s ID** | **Patient’s age range** | **Patient’s Gender** | **Tumor Fraction** | **Sturgeon Classification** | **Sturgeon Confidence Score** | **Pathology and Other Molecular Method Classification** | **Pathology and Other Molecular Method Used** |
| --- | --- | --- | --- | --- | --- | --- | --- |
| Sch.1 | 0-5Y | f | 90 | methylation class atypical teratoid/rhabdoid tumor, subclass MYC | 0.99 | Atypical teratoid rhabdoid tumor, WHO grade 4 | Pathology, Illumina Methylation EPIC Array |
| Sch.2 | 0-5y | m | 90 | methylation class low grade glioma, subclass posterior fossa pilocytic astrocytoma | 0.97 | Low grade glioma (LGG) compatible with Pilocytic astrocytoma, WHO grade 1 | Pathology, NGS |
| Sch.3 | 0-5Y | m | 95 | methylation class meningioma | 0.97 | Atypical meningioma, WHO grade 2 | Pathology |
| Sch.4 | 0-5Y | m | 100 | methylation class medulloblastoma, subclass group 3 | 0.97 | Medulloblastoma, WHO grade 4, non-WNT/non-SHH subgroup. | Pathology, Nanostring |
| Sch.5 | 6-10Y | m | 95 | Diffuse midline gliomas, H3K27-altered | 0.95 | Midline pontine diffuse glioma like | Pathology, NGS |
| Sch.6 | 0-5Y | m | 80 | methylation class ependymoma, Posterior Fossa A group | 0.93 | Ependymoma, Posterior fossa-A group,WHO grade 2 | Pathology |
| Sch.7 | 6-10Y | m | 90 | methylation class low grade glioma, subclass posterior fossa pilocytic astrocytoma | 0.93 | Low grade glioma (LGG) compatible with Pilocytic astrocytoma, WHO grade 1 | Pathology, Oncomine |
| Sch.8 | 6-10Y | f | 40 | methylation class embryonal tumor with multilayered rosettes | 0.9 | Malignant embryonal tumor suspicious for embryonal tumor with multilayered rosettes, WHO grade 4 | Pathology,Illumina Methylation EPIC Array NGS, Oncomine |
| Sch.9 | 0-5Y | f | 20 | methylation class atypical teratoid/rhabdoid tumor, subclass MYC | 0.9 | Atypical teratoid rhabdoid tumor, WHO grade 4 | Pathology, SNP Array,NGS, Oncomine |
| Sch.10 | 0-5Y | f | unavailable | methylation class CNS high grade neuroepithelial tumor with BCOR alteration | 0.9 | CNS high grade neuroepithelial tumor with BCOR alteration | Pathology, Foundation One |
| Sch.11 | 16-20Y | f | 30 | methylation class melanotic schwannoma | 0.8 | Schwannoma showing areas of an increased proliferation index | Pathology, NGS, |
| Sch.12 | 6-10Y | f | 80 | methylation class control tissue, hypothalamus | 0.8 | LGG-with features of pilocytic astrocytoma | Pathology, NGS |
| Sch.13 | 11-15Y | f | 100 | methylation class low grade glioma, subclass posterior fossa **pilocytic astrocytoma** | 0.7 | LGG-with feature of **pilocytic astrocytoma** | Pathology, NGS |

***SI Table 3: Validation cohort***

*Each row corresponds to a sample in the cohort. In addition, columns display age and gender, Sturgeon classification result and score, classification obtained by other established methods, and method type.*

Below are additional radiograms and histology images for the case studies discussed in the main text.

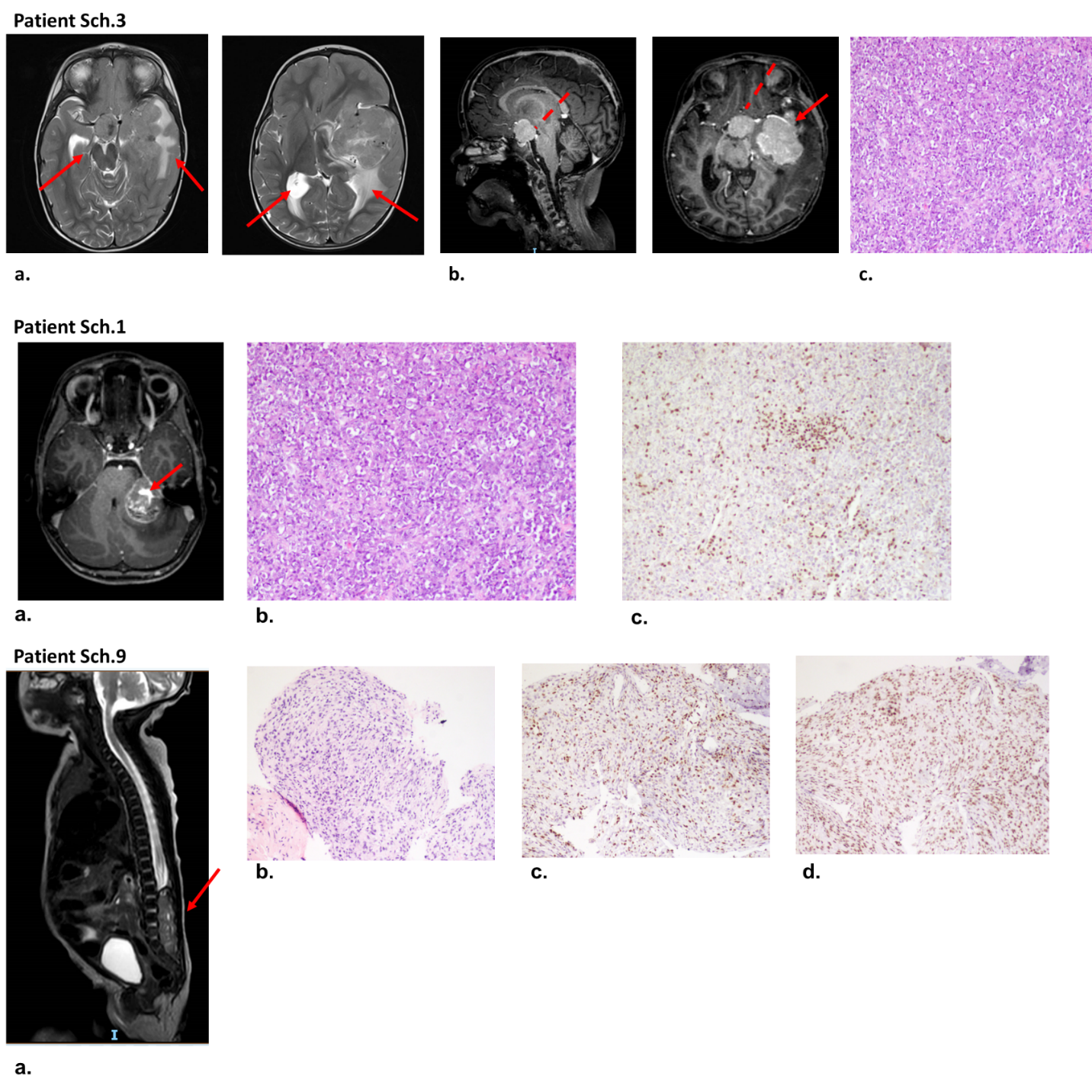

***SI Figure 4: Clinical data of case study patients***

***Patient Sch.3. a.*** *MRI revealed a substantial left parieto-temporal tumour with suspected leptomeningeal spread.* ***b.*** *T1 axial and sagittal images with Gad. Full line = left temporal tumor. Dashed line suprasellar tumor. There is mass effect on the midbrain. They both show homogeneous enhancement.*

***c.*** *H&E staining of Atypical teratoid rhabdoid tumor, WHO grade 4. Picture showing sheets of densely packed atypical cells with rhabdoid features.*

***Patient Sch.1: . a.*** *MRI scan shows a tumor in the left cerebellopontine angle with displacement of the 4^th^ ventricle, T1 axial with Gadolinium. The tumor shows heterogeneous enhancement.*

***b.*** *H&E staining****. c.*** *Histochemical staining done by pathologist, for biopsy from the tumor strongly suggested ATRT, but staining for INI1 (SMARCB1) essential for this classification, showed a mosaic pattern.*

**FFPE embedding protocol**

The FFPE tissues in this study were processed with standard methods, briefly: Initially the tissues are stored in formalin right after their removal by the surgeon. Tissue typically spend 1-5 days in formalin before cutting of tumour region as identified by a pathologist. Following, the tissues are processed automatically by Tissue-Tek VIP 6 (MODEL VIP 6-E2). This is followed by embedment in paraffin (wax) to form blocks.  Finally, the blocks are cut to 4-5 micrometre (μm) unstained sections and placed on slides for staining.
